# Dengue antibodies can cross-react with SARS-CoV-2 and *vice versa*-Antibody detection kits can give false-positive results for both viruses in regions where both COVID-19 and Dengue co-exist

**DOI:** 10.1101/2020.07.03.20145797

**Authors:** Himadri Nath, Abinash Mallick, Subrata Roy, Soumi Sukla, Keya Basu, Abhishek De, Subhajit Biswas

## Abstract

Five of thirteen Dengue antibody-positive serum samples, dated 2017 (pre-dating the COVID-19 outbreak) produced false-positive results in SARS-CoV-2 IgG/IgM rapid strip tests. Our results emphasize the importance of NAT and/or virus antigen tests to complement sero-surveillance for definitive diagnosis of COVID-19/Dengue in regions where both viruses are co-endemic.

## Introduction

The world is experiencing the COVID-19 pandemic with 9,843,073 confirmed cases and 4,59,760 deaths till 28^th^ June, 2020(1). SARS-CoV-2 infection is increasing in India with about 18-19 thousand confirmed cases being reported daily for last few days (1). Due to this high daily infection rate, rapid tests for COVID-19 antibodies (Abs) are being increasingly implemented to detect onset of community transmission, if any, especially the asymptomatic and convalescent cases.

It has been already reported from Singapore that Abs elicited by SARS-CoV-2 infection can produce false-positive results in Dengue IgG and IgM rapid tests (2). It is also noteworthy that early symptoms of COVID-19 can be mistaken for Dengue fever including thrombocytopenia in highly dengue endemic countries like India and Brazil (3).

By this time, with the onset of monsoon in India, Dengue infections have started increasing with COVID-19 pandemic in the background. In this scenario, the obvious question is whether DV Abs, prevalent in people in highly Dengue endemic regions, will cross-react in COVID-19 rapid tests. If this happens, serology-based diagnosis and sero-surveillance for these immunologically cross-reacting viruses have to be carried out with adequate precautions/background and other supporting information, in regions where both viruses are co-existent. Interpretation of results has to be done with care to avoid arriving at erroneous conclusions.

## Result & Discussion

Five of the thirteen DV Ab-positive samples were found to produce false-positive bands in SARS-CoV-2 IgG and IgM rapid test. Same DV Ab-positive sample was found to produce false-positive result in two different COVID-19 test kits (Figure 1). This confirms that DV Abs can, indeed, cross-react with SARS-CoV-2 antigen(s) and give false-positive results in COVID-19 rapid IgG and IgM test (Table 1).

**Table 1:** Rapid IgG and IgM test results for COVID-19 and Dengue*

| Sample Name | Dengue IgG | Dengue IgM | COVID IgG | COVID IgM |
| --- | --- | --- | --- | --- |
| 17-D-59 | f+ | +++ | - | ++ |
| 17-D-68 | + | - | - | - |
| 17-D-12 | ++ | + | - | + |
| 17-D-1 | + | + | - | - |
| 17-D-7 | ++ | - | - | + |
| 17-D-11 | + | + | - | - |
| 17-D-25 | ++ | - | - | + |
| 17-D-31 | + | - | - | - |
| 17-D-30 | + | - | - | - |
| 17-D-48 | ++ | + | - | - |
| 17-D-37 | ++ | - | - | - |
| 17-D-15 | + | ++ | ++ | - |
| 17-D-50 | - | ++ | - | - |
| 17-D-16 (NC) | - | - | - | - |

\* “+” sign denotes positive result; number of “+” sign signifies relative increase in positive
band intensity in the strip tests. “f+” stands for faint-positive.
“-” sign signifies negative result.
NC stands for negative control.

**Figure 1:**
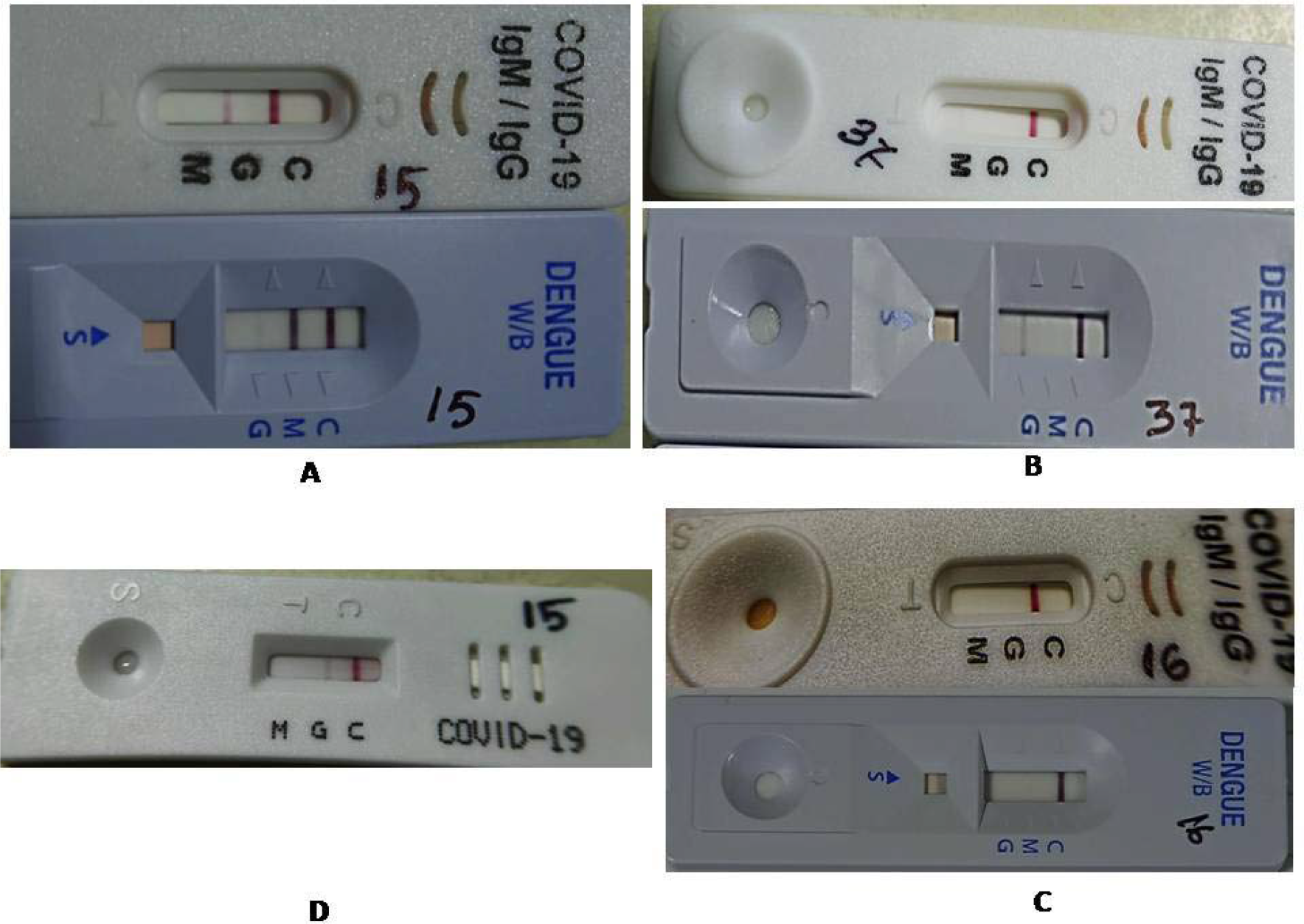
Representative images of COVID-19 and Dengue rapid IgG and IgM strip tests (A) Sample 17-D-15; (B) Sample 17-D-37; (C) Sample 17-D-16; (D) Sample 17-D-15 test using AbCheck, kit. A, D, represent a serum sample, confirmed positive for Dengue but false-positive for COVID-19. B, represents a serum sample, confirmed positive for Dengue but negative for COVID-19. C, is a serum sample negative for both Dengue and COVID-19 antibodies.

The aforesaid antibody test results, in fact, confirmed our computational modelling (docking) studies that predicted with high confidence that human antibodies to anti-DV envelope can potentially bind to “receptor-binding motif (RBM)” of SARS-CoV-2 Spike protein with some of the interactions even intercepting the ACE2 receptor binding to RBM (4). As COVID-19 rapid Ab test kits mostly use immobilized SARS-CoV-2 surface antigen(s), our prediction is supported by the observed DV false-positivity in COVID-19 Ab tests.

Our results demonstrate that in Dengue endemic countries, Ab detection-based assays can result in false-positive reports for COVID-19 for actually DV-infected patients. The reverse scenario has been reported from Singapore, i.e. originally COVID-19 patients were mis-diagnosed as having Dengue as antibodies to SARS-CoV-2 cross-reacted in Dengue antibody tests (2). From the above scenarios and our computational modelling studies, it appears that both these viruses have some antigenic similarity that is resulting in the observed cross-reactivity and warrants further investigation to elucidate this.

So, sero-surveillance needs to be complemented with NAT and/or virus antigen testsfor definitive diagnosis of COVID-19 and Dengue in regions where both the viral diseases are co-existent (5).

## Methods

We have performed rapid DV IgG and IgM tests (SD Bioline, Abbott) on archived serum samples from DV-diagnosed patients (NS1 ELISA-positive) from the 2017 Dengue outbreak in Kolkata (pre-dating COVID-19 pandemic). Thirteen DV-Ab positive samples were then subjected to rapid SARS-CoV-2 IgG and IgM strip test (ImmunoQuick, ImmunoScience India) following manufacturers’ instructions. AbCheck COVID-19 IgG and IgM test kit (NuLifecare) was also used to confirm the cross-reactivity (Figure 1D).

In brief, 20ul of each sample was added in specified area of test strips followed by addition of two drops (∼80-100ul) of kit-specific assay buffer in the designated spot, depending on the test kit. Assay buffer was added in marked region in case of Dengue Ab kit and for COVID-19 rapid kits, at the same position where sample was added first. Appearance of “test line” for all strip testswas confirmed to ensure the validity of the tests performed. We have also used negative control serum (both Dengue and COVID-19 negative) as shown (Figure 1C).

## Data Availability

All data are available from the corresponding author on request.

## Acknowledgements

The authors acknowledge CSIR-IICB for providing laboratory facilities for conducting the present work.

## Financial support

The project was funded by a grant from the **Council of Scientific and Industrial Research, India** to S.B. Grant number: MLP 130; CSIR Digital Surveillance Vertical for COVID-19 mitigation in India.

## Potential conflict of interest

The authors declare no competing financial interests.

## Ethical approval and informed consent

This study was performed in accordance with the ethical standards (at par with the 1964 Helsinki declaration and its later amendments) of the review boards of all relevant institutions. Informed consent was obtained from all individual participants included in the original study. All experiments were carried out in accordance with the relevant guidelines and regulations.

## Notes

### Competing Interest Statement

The authors have declared no competing interest.

### Author Declarations

Ethical approval for the research was granted by the respective Institutional Ethical Committee of CSIR-IICB and Calcutta National Medical College, Kolkata.

